# A longitudinal evaluation of personalized intrinsic network topography and cognitive decline in Parkinson’s disease

**DOI:** 10.1101/2023.02.03.23285447

**Authors:** Renxi Li, Vincent Pozorski, Kevin Dabbs, Maureen Haebig, Rasmus Birn, Colleen Pletcher, Alexandra Wey, Amy Barzgari, Frances Theisen, Christopher Cox, Ozioma Okonkwo, Catherine Gallagher

**Affiliations:** William S. Middleton V.A. Hospital, Madison, WI, USA; Department of Neurology, University of Wisconsin School of Medicine and Public Health, Madison, WI, USA; Department of Psychiatry, University of Wisconsin School of Medicine and Public Health, Madison, WI, USA; Louisiana State University, Department of Psychology, Baton Rouge, LA, USA; Department of Medicine, University of Wisconsin School of Medicine and Public Health, Madison, WI, USA; Wisconsin Alzheimer Disease Research Center, Madison, WI, USA

**Author notes:** Corresponding Author: Catherine L. Gallagher, MD; 2500 Overlook Terrace Dr., Room B6053, Madison, WI. 53705;. Li: The George Washington University School of Medicine and Health Sciences, Washington, DC, USA Wey: Medical College of Wisconsin, Milwaukee, WI, USA. Barzgari: University of Illinois School of Medicine, Rockfield, USA.

**Keywords:** humans, aged, disease, prognostic markers, cerebral cortex, brain connectivity, network analysis, dementia

## Abstract

Resting state functional MRI (R-fMRI) offers insight into how synchrony within and between brain networks is altered in disease states. Individual and disease-related variability in intrinsic connectivity networks may influence our interpretation of R-fMRI data.

**Methods:** We used a personalized approach designed to account for individual variation in the spatial location of correlation maxima to evaluate R-fMRI differences between Parkinson’s disease (PD) patients who showed cognitive decline, those who remained cognitively stable, and cognitively stable controls. We compared fMRI data from these participant groups, studied at baseline and 18 months later, using both Network-based Statistics (NBS) and calculations of mean inter- and intra-network connectivity within pre-defined functional networks.

**Results:** The NBS analysis showed that PD participants who remained cognitively stable showed exclusively (at baseline) or predominantly (at follow-up) increased intra-network connectivity, whereas decliners showed exclusively reduced intra-network and inter-(ventral attention and default mode) connectivity, in comparison to the control group. Evaluation of mean connectivity between all ROIs within a priori networks showed that decliners had consistently reduced inter-network connectivity for ventral attention, somatomotor, visual, and striatal networks, and reduced intra-network connectivity for ventral attention network to striatum and cerebellum.

**Conclusions:** These findings suggest that specific functional connectivity covariance patterns differentiate PD cognitive subtypes and may predict cognitive decline. Further, increased intra and internetwork synchrony may support cognitive function in the face of PD-related network disruptions.

**Key Points:**

- Resting state functional MRI (R-fMRI) can be used to probe changes in brain networks related to disease. Personalized approaches can be used to address spatial variations in R-fMRI correlation maxima influenced by individual variation or brain plasticity in response to disease.
- Longitudinal R-fMRI from cognitively stable Parkinson’s patients were compared with those who experienced decline as well as controls using a personalized approach.
- Cognitively stable patients showed increased inter and intra-network synchrony while decliners showed decreases that may have preceded cognitive decline.

## Introduction

Cognitive decline is a frequent, if not inevitable, non-motor complication of Parkinson’s disease (PD). Since the temporal onset and domain-specific impairment varies between individuals and PD subtypes, it would be useful to have a non-invasive biomarker that predicts decline. As both cognitive and motor symptoms of PD are related to dysregulation of frontal-subcortical and frontal-cerebellar circuits, resting state functional MRI (R-fMRI), which can evaluate the degree of blood oxygen dependent (BOLD) synchrony between multiple brain regions at once, has appeal as a biomarker.

While R-fMRI is a useful tool in clinical research, its replicability has faced challenges, particularly in disease states (Griffanti et al., 2016; Zuo et al., 2010). In typical MRI processing pipelines, the brain is mapped into a standard space or surface, based on anatomical characteristics. These approaches assume that the same anatomical brain area in each individual is responsible for the same function. However, recent studies have shown spatial variability in functional networks between individuals, especially within heteromodal cortices (Mueller et al., 2013). Furthermore, brain diseases are often accompanied by plastic adaptations in cortical function that likely impact network architecture (Calabresi et al., 2007; Nudo, 2007). Personalized approaches, in which the anatomic locations of connectivity maxima are allowed to vary slightly between individuals, have been introduced as a way of addressing this problem (Dickie et al., 2018).

For this study, we investigated longitudinal R-fMRI differences between non-demented PD patients who remained cognitively stable (CSPD), those who showed cognitive decline (CDPD), and stable controls, studied at baseline and 18 months. Following standard R-fMRI preprocessing and denoising, we applied Personalized Intrinsic Network Topography (PINT) (Dickie et al., 2018), a novel approach first developed for use in autism, to individualize the locations of key network nodes. Using this approach, we found differences in the strength of inter- and intra-network correlations, many of which were stable across visits.

## Methods

### Participants, demographics, and characteristics

A total of 85 participants were recruited through local neurology clinics and the Wisconsin Alzheimer’s Disease Research Center as part of the VA-sponsored “Longitudinal MRI in Parkinson’s Disease (LMPD)” study. Participants were asked to complete two visits 18 months apart, providing written informed consent at the first study visit prior to research procedures. Exclusion criteria included known cardiovascular and cerebrovascular disease, dementia or mild cognitive impairment (MCI), major psychiatric or neurological disease other than PD, and baseline Mini-Mental State Examination (MMSE) (Folstein et al., 1975) score of less than 27/30, and for PD participants, symptom onset earlier than age 45. Baseline and follow-up visits each consisted of a neuropsychological assessment, Unified Parkinson’s Disease Rating Scale (UPDRS) (Fahn, S. & Elton, R., 1987) scoring by a movement disorders neurologist (C. G.), and brain MRI. PD participants were off anti-Parkinson medications for 12-18 hours prior to each visit. The study was approved by the University of Wisconsin Health Sciences IRB and endorsed by the William S. Middleton VA’s R&D committee.

### Neuropsychological assessment and cognitive subtype assignment

To quantify participants’ performance in cognitive domains of executive function, memory, language, and baseline literacy, a series of tests was administered at both baseline (v1) and follow-up (v2) visits, using different versions of the tests when available. Tests administered included (1) Boston Naming Test (BNT) (Kaplan et al., 1983), (2) category (Goodglass & Kaplan, 1972; Rosen, 1980) and phonemic fluency tests (C-F-L form) (Benton & Hamsher, 1976), (3) Hopkins Verbal Learning Test (HVLT) (Benedict et al., 1998; Brandt, 1991), (4) Trail Making Tests (TMT) A and B (Reitan, 1992), (5) Wide Range Achievement Test - Word Reading Subset Score Fourth Edition (WRAT-IV) reading test (Wilkinson & Robertson, 2006), and (6) Wisconsin Card Sort Test - 64 (WCST-64) (Grant & Berg, 1948; Greve, 2001). For participants, z-scores based on the control group mean and SD were generated for each time point. These z-scores were combined into three composite measures as follows:

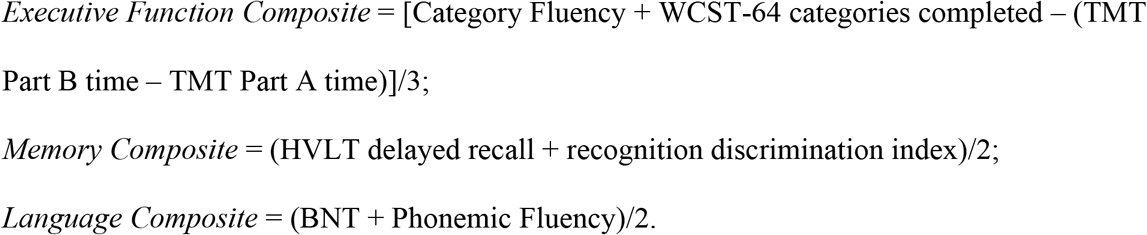

To define the cognitive subtypes (stable versus decline over the 18-month interval), each participant’s z-score at baseline (v1) was subtracted from the follow-up (v2) score to calculate a change score. Participants who experienced 1 or more SD decline in two or more (of memory, language, and executive) domains between baseline and follow-up were defined as decliners, with the remainder being cognitive stable.

Statistical analysis of group demographic information and disease characteristics was carried out using IBM SPSS (version 23.0). Comparisons of non-dichotomous variables, such as age and education, were performed using two-tailed independent sample t-tests as well as Analysis of Variance (ANOVA) followed by Tukey-Kramer post-hoc comparison. Dichotomous variables, such as sex, were evaluated using the Fisher-Freeman-Halton test(Freeman & Halton, 1951). For baseline neuropsychological data, between-group comparisons were derived via Analysis of Covariance (ANCOVA), with age, sex, and WRAT-IV as covariates. When comparing the longitudinal (v2-v1) scores, age, sex, WRAT-IV, and time between visits were controlled in the ANCOVAs. Benjamini-Hochberg procedure at *α* = 0.05 was used for false discovery rate (FDR) correction following the ANCOVAs. The Bonferroni method (p < 0.05/3) was used to correct for multiple comparisons in the post hoc pairwise tests.

### MRI acquisition

Brain MRIs were acquired using a GE 750 Discovery 3T MRI system (General Electric Healthcare, Waukesha, WI) with an eight-channel phased array head coil. A high-resolution 3D brain volumetric (BRAVO) T1-weighted inversion prepared sequence of inversion time (TI) = 450 ms, repetition time (TR) = 8.2 ms, echo time (TE) = 3.2 ms, flip angle = 12°, acquisition matrix = 256 × 256, field of view (FOV) = 256 mm, and slice thickness = 1.0 mm, collected in the axial plane, was obtained from each participant as structural data for the segmentation of gray matter (GM), white matter (WM), and cerebrospinal fluid (CSF). R-fMRI data were acquired using a gradient recalled echo type echo-planar imaging (GRE-EPI) imaging sequence of TR = 2000 ms, flip angle = 70°, acquisition matrix = 64 × 64, and FOV = 240 mm. Thirty-six contiguous 4 mm slices were acquired every 2 seconds over 7 minutes for a total of 210 volumes of images with a voxel size of 3.75 mm x 3.75 mm x 4 mm.

### Image preprocessing

Following visual inspection for artifacts, subject-specific resting-state BOLD images were coregistered to their respective T1-weighted images using the Statistical Parametric Mapping (SPM12; https://www.fil.ion.ucl.ac.uk), implemented in MATLAB (MathWorks, Inc., Natick, MA). Structural and functional data were preprocessed according to the Functional Connectivity Toolbox (CONN version 18b) (Whitfield-Gabrieli & Nieto-Castanon, 2012) default preprocessing pipeline, which included removal of the first 3 functional volumes, realignment, slice-time correction, outlier identification using the Artifact Detection Tool (ART; https://www.nitrc.org/projects/artifact_detect), tissue segmentation, normalization to Montreal Neurologic Institute (MNI) space, resampled at 2 mm x 2 mm x 2 mm, and smoothing with a Gaussian kernel of 8 mm^3^ full-width half-maximum.

To restrict the analyses to gray matter, T1-based subject-specific implicit masks (based on an 80% threshold on the global T1 signal) were applied to the functional images. These resting-state time courses were then corrected for residual physiological and motion artifacts according to the default denoising steps in CONN, which included the removal of invalid volumes identified during ART-based outlier detection, and regression-based removal of 10 principal components of WM and CSF (CompCor) (Behzadi et al., 2007), 6 motion parameters, and their 6 first-order temporal components defined during the realignment preprocessing step. Finally, BOLD time course data were bandpass filtered to include 0.008 and 0.09 Hz frequencies representative of slow neuronal fluctuations and converted to percent signal change.

### Surface-based registration of the functional data

Automatic segmentation of each participant’s T1-weighted image from the baseline visit was carried out in the FreeSurfer (v6.0.0;(Fischl, 2012)), followed by inspection and revision of the grey-white matter and pial surfaces as necessary (A.W., F.T., A.B., and C.P.) Inter-rater reliability Spearman’s correlation for Freesurfer preprocessing in our lab is 0.98. The default ciftify anatomical pipeline (Dickie et al., 2019) was then applied to each subject’s FreeSurfer reconstruction to enable precise surface-based registration of the cerebral cortex. Here, a 2D coordinate system based on topographical properties (e.g. segmentation of sulci and gyri), rather than the absolute positions in 3D space, was used to represent cortical anatomical structures. Lastly, the preprocessed functional images from CONN were processed using the default ciftify functional pipeline (Dickie et al., 2019) where fMRI volumes were mapped to subject-specific gray coordinate surfaces in a weighted and ribbon-constrained way.

### Region of interest (ROI) definition

PINT enables the identification of correlation maxima within predefined cortical networks, allowing for slight variation in precise anatomical locations of functional maxima between individuals. These adjustments were made to 80 template 6 mm regions of interest (ROIs) in 6 resting-state cortical networks from a seven-network atlas (Yeo et al., 2011), which included dorsal attention (12 ROIs), default mode (14 ROIs), ventral attention (18 ROIs), frontoparietal (16 ROIs), sensory motor (10 ROIs), and visual (10 ROIs) networks as shown in Figure 1. No ROI was chosen to present the limbic network due to its high fMRI signal susceptibility (Dickie et al., 2018).

**Figure 1.**
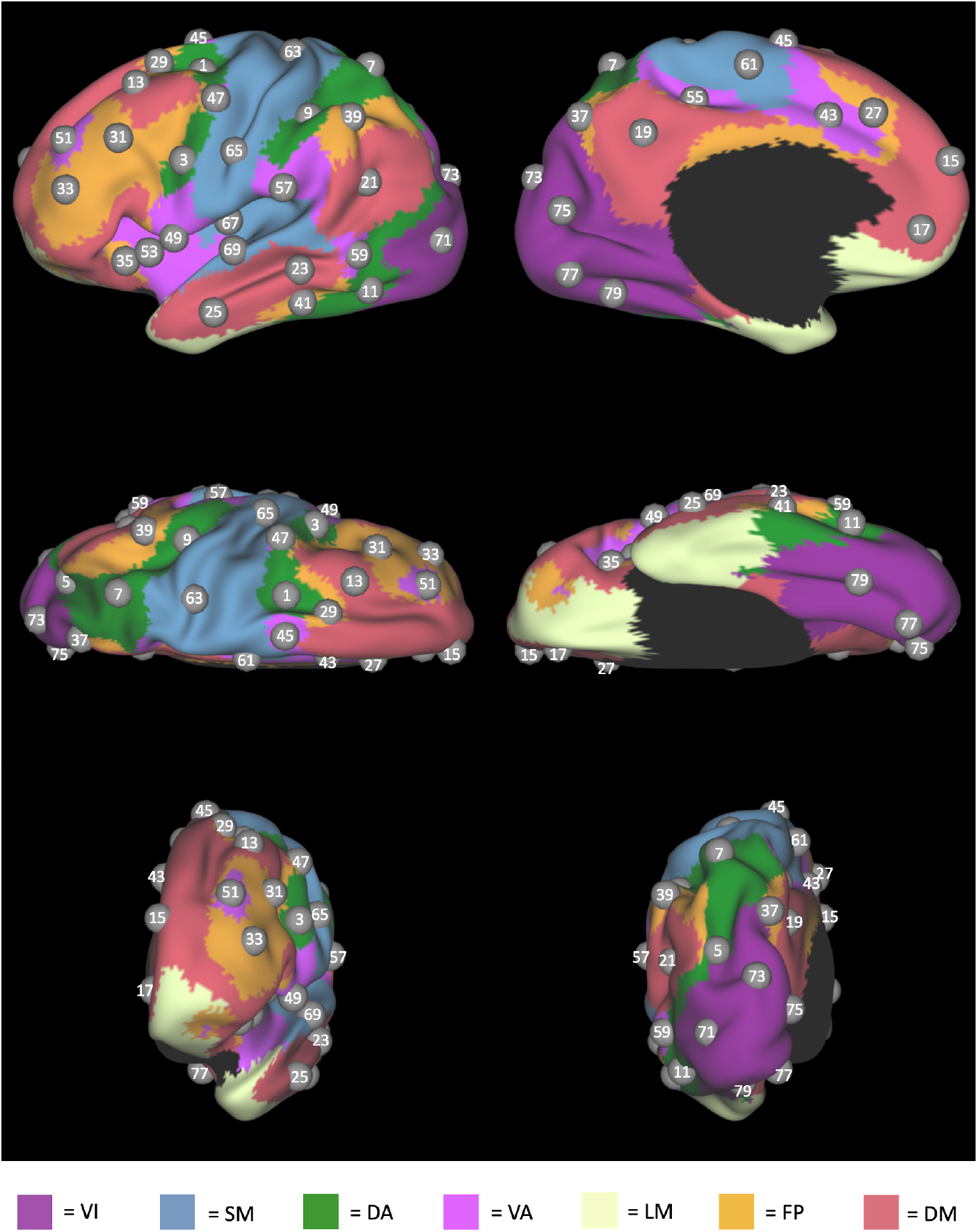
Regions of Interest. Eighty regions of interest (ROIs), shown on the left cerebral surface, defined based on Yeo’s seven-network atlas prior to the application of PINT. Corresponding right hemisphere ROI labels are one integer greater than the left hemisphere. ***Abbreviations:*** DA, dorsal attention; DM, default mode; FP, frontoparietal; LM, limbic; SM, somatomotor; VA, ventral attention; VI, visual.

The before- and after-PINT correlation maps for the eighty cortical ROIs in an example participant are shown in Figure 2. The intra-network connectivity, represented by the groups of vertices along the correlation map diagonal, increases (as expected) after application of the PINT algorithm. Besides visual quality checks from the correlation maps for each participant, the reliability of PINT on this dataset was tested by comparing the longitudinal (v2-v1) within-versus between-subject ROI-shift distances (Dickie et al., 2018) The within-subject ROI-shift distance (4.41 ± 4.11 mm; n = 5680) was significantly smaller (p < 0.001) than its between-subject counterpart (6.21 ± 5.43 mm; n = 403280).

**Figure 2.**
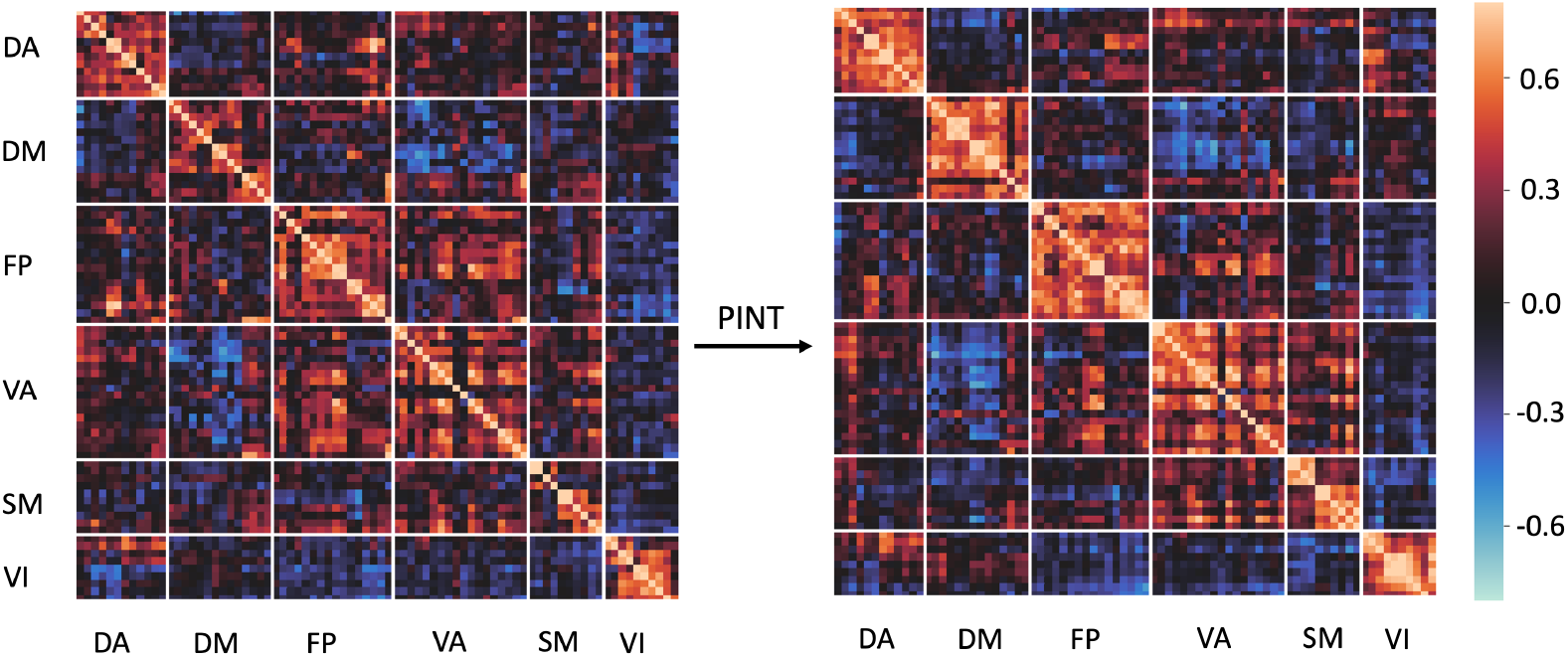
Application of Personalized Intrinsic Network Topography (PINT) in an example participant, visualized via a correlation matrix. PINT adjusts the spatial locations of eighty cortical template ROIs, shown in Figure 1, within a 6-mm search radius to maximize intra-network partial correlations. ***Abbreviations:*** DA, dorsal attention; DM, default mode; FP, frontoparietal; LM, limbic; SM, somatomotor; PINT, Personalized Intrinsic Network Topography; VA, ventral attention; VI, visual.

Because PINT does not presently incorporate subcortical and cerebellar functional networks, previous functional mapping of the cerebellum (Buckner et al., 2011) and striatum (Choi et al., 2012) were used to define 14 cerebellar and 14 striatal ROIs. Definition of the subcortical ROIs was based on their functional connectivity to Yeo’s seven-network atlas (Yeo et al., 2011) for the 80 cortical ROI template used in the PINT (i.e. each subcortical ROI was functionally connected to a proposed cerebral network). The resulting cerebellar and the striatal ROIs specific to our data are shown in Supplemental Figures 1 and 2, respectively.

### Network-Based Statistic (NBS) analysis

For each participant, the resting-state time courses of the 80 PINT-individualized cortical ROIs and 28 subcortical ROIs were extracted and used to calculate a subject-specific correlation matrix for the baseline (v1) as well as the follow-up (v2) time points. A longitudinal (v2-v1) subject-specific matrix was calculated by subtracting the baseline (v1) matrix from follow-up matrix (v2). The Fisher-z transformation was applied to all correlation coefficients.

Using the baseline (v1), follow-up (v2), and longitudinal (v2-v1) subject-specific correlation matrices, Network-Based Statistic (Zalesky et al., 2010) analyses were used to compare the strength of each ROI-ROI pair connection between participant groups using 2000 permutations. Since we found age to be significantly correlated with mean intra-/inter-network functional connectivity between and within several networks, baseline age was included as a covariate in all NBS ANCOVAs. NBS results were False Discovery Rate (FDR)-corrected with p-corrected < 0.05 as the significance level.

### Mean intra- and inter-network comparison

Since both intra and inter-network ROI-ROI correlations showed high functional dependency, the NBS investigation of ROI-ROI connectivity might be limited if a network-wide difference existed without significant ROI-ROI differences. To investigate overall connectivity differences between the PD cognitive groups and stable controls at baseline (v1) and follow-up (v2), the control group ROI-ROI connectivity matrix at baseline was subtracted from the CSPD or CDPD counterpart. Likewise, CDPD was subtracted from CSPD. ROI-ROI connectivity differences within and between networks were then averaged across ROIs within the pre-defined functional networks (dorsal attention, DA; default-mode, DM; frontoparietal, FP; ventral attention, VA; somatomotor, SM, visual, VI; cerebellum, CRB; and striatal, STR). To account for the non-independency of ROI-ROI correlations, covariances between all ROIs in a given ROI-ROI comparison were calculated to derive the uncertainty of the intra-or inter-network connectivity. Longitudinal (v2-v1) comparisons followed the same procedure using the respective group ROI-ROI connectivity differences between visits and calculated covariances.

The effect size was calculated from Welch’s t-test to compare each mean intra-or inter-network connectivity difference between CSPD and CSPD-control. Significance was determined using Benjamini-Hochberg FDR correction for multiple comparisons at a 0.05 level. Then, due to imperfect age matching between CDPD and the comparison groups, comparison of differences in mean network connectivity between CSPD, CDPD, and age-matched subgroups of control participants were conducted using the same methods.

## Results

Of the original 85 enrollees, 71 (41 controls and 30 PD) completed both baseline and follow-up visits; one control participant did not have sufficient f-MRI data for analysis. Thirty-five control and 20 PD participants remained cognitively stable between visits 1 and 2, while 5 control and 10 PD participants experienced decline. Therefore, final participants’ data analyzed for this study were those from 35 cognitively stable controls, 20 CSPD participants, and 10 CDPD participants. Group characteristics at baseline (v1) are summarized in **Table 1**. The CDPD group (73.7 ± 6.8 yrs) was older than both the control (66.0 ± 7.4 yrs) and the CSPD (62.6 ± 8.8 yrs) groups. Both the CSPD (29.1 ± 1.0) and CDPD (29.1 ± 1.0) had lower MMSE scores than the control (29.7 ± 0.6). While all three groups were predominantly male, the ratios of men to women did not differ between them. Although years of formal education were fewer in CSPD (15.5 ± 2.4 yrs) than control (17.5 ± 2.4 yrs), WRAT-IV scores, an estimate of quality of education, did not differ between groups.

**Table 1.** Participant characteristics at baseline

|  | **Control**<br>**(n = 35)** | **CSPD**<br>**(n = 20)** | **CDPD**<br>**(n = 10)** | **P-value** |
| --- | --- | --- | --- | --- |
| Age at enrollment (y) | 66.0 ± 7.4 | 62.6 ± 8.8 | 73.7 ± 6.8 | 0.002<sup>††, b, c</sup> |
| Sex (Male/Female) | 25/10 | 16/4 | 8/2 | 0.730<sup>†</sup> |
| Education (y) | 17.5 ± 2.4 | 15.5 ± 2.4 | 16.9 ± 3.4 | 0.024<sup>††, a</sup> |
| WRAT-IV | 62.4 ± 3.8 | 62.0 ± 5.5 | 61.4 ± 4.9 | 0.831 |
| MMSE | 29.7 ± 0.6 | 29.1 ± 1.0 | 28.7 ± 0.8 | 0.001<sup>††, a, b</sup> |
| Age at diagnosis (y) | N/A | 59.2 ± 8.7 | 69.1 ± 6.2 | 0.001 |
| Disease duration (y) | N/A | 3.4 ± 3.2 | 4.6 ± 3.4 | 0.396 |
| Side of onset (R/L/S) | N/A | (12/7/1) | (4/3/3) | 0.229<sup>†</sup> |
| Hoehn & Yahr Stage | N/A | 1.7 ± 0.7 | 1.6 ± 0.6 | 0.688 |
| UPDRS III | N/A | 20.7 ± 12.1 | 20.4 ± 8.4 | 0.938 |
| UPDRS Total | N/A | 35.6 ± 15.3 | 33.9 ± 12.1 | 0.744 |

**Legend:** Baseline values (mean ± SD) are shown.
P-values for between-group comparisons were derived via two-tailed independent samples t-test, <sup>†</sup>Fisher-Freeman-Halton test, and <sup>††</sup>ANOVA followed by Tukey-Kramer post-hoc comparison with significant group differences as follows:
<sup>a</sup> CSPD versus controls; <sup>b</sup> CDPD versus controls; <sup>c</sup> CSPD versus CDPD.
**Abbreviations:** ANOVA, one-way analysis of variance; CDPD, cognitive decline Parkinson's disease; CSPD, cognitively stable Parkinson's disease; L, left; MMSE, Mini-Mental State Examination; N/A, not applicable; R, right; S, symmetrical; SD, standard deviation; UPDRS, Unified Parkinson's Disease Rating Scale; WRAT-IV, Wide Range Achievement Test - Word Reading Subset Score Fourth Edition.

In regard to Parkinson’s disease, the CDPD group was diagnosed with PD at a later age (69.1 ± 6.2 years) than CSPD patients (59.2 ± 8.7 yrs). Disease duration and laterality of symptom onset were not different between CSPD (3.4 ± 3.2 yrs; Right 12/Left 7/Symmetrical 1) and CDPD (4.6 ± 3.4 yrs; Right 4/Left 3/Symmetrical 3) patients. At the time of the baseline study visit, the CSPD patients had an average Hoehn and Yahr stage of 1.7 ± 0.7, a UPDRS III (motor) subscore of 20.7 ± 12.1, and a total UPDRS score of 35.6 ± 15.3 (Fahn & Elton, 1987). None of these scores were significantly different from the CDPD patients, whose average Hoehn and Yahr stage was 1.6 ± 0.6, UPDRS III subscore was 20.4 ± 8.4, and UPDRS total score was 33.9 ± 12.1.

Neuropsychological composite z-scores at baseline (v1) and rates of change (v2-v1) in these composites over the 18-month interval are summarized in **Table 2**. At baseline (v1), both the CSPD and the CDPD patients had lower executive function composite z-scores than stable controls. In addition, the CDPD group had a lower baseline mean executive composite z-score (−1.2 ± 1.2) than the CSPD group (−0.34 ± 0.91). However, age was correlated with executive composite scores in both PD and control groups (p = 0.005).

**Table 2.** Baseline composite z-scores and longitudinal differences

|  | **Control**<br>**(n = 35)** | **CSPD**<br>**(n = 20)** | **CDPD**<br>**(n = 10)** | **F, P-value** |
| --- | --- | --- | --- | --- |
| Executive function composite *z-score*<sub>v1</sub> | 0.07 ± 0.12 | -0.48 ± 0.16 | -0.75 ± 0.24 | 7.93, 0.001<sup>a, b</sup> |
| Executive function composite *z-score*<sub>v2-v1</sub> | -0.02 ± 0.10 | -0.02 ± 0.14 | -0.95 ± 0.20 | 8.78, < 0.001<sup>b, c</sup> |
| Memory composite *z-score*<sub>v1</sub> | 0.02 ± 0.20 | -0.73 ± 0.27 | -0.91 ± 0.40 | 4.09, 0.022 |
| Memory composite *z-score*<sub>v2-v1</sub> | 0.35 ± 0.13 | 0.09 ± 0.17 | -0.59 ± 0.25 | 5.54, 0.006<sup>b</sup> |
| Language composite *z-score*<sub>v1</sub> | 0.06 ± 0.13 | -0.03 ± 0.18 | -0.30 ± 0.26 | 0.76, 0.471 |
| Language composite *z-score*<sub>v2-v1</sub> | 0.21 ± 0.13 | 0.23 ± 0.17 | -0.11 ± 0.25 | 0.70, 0.499 |

**Legend:** Composite z-scores are shown as mean ± SD.
Following multiple comparisons correction, between-group differences are indicated as follows:
<sup>a</sup> CSPD versus controls; <sup>b</sup> CDPD versus controls; <sup>c</sup> CSPD versus CDPD.

### NBS Results

Significant between-group ROI-ROI connectivity differences are visualized as cord diagrams in Figure 3 with specific ROI’s listed in Tables 3-5. Odd numbered ROIs are located in the left hemisphere and corresponding even numbered ROIs of one integer greater in the right hemisphere. Compared to the control group, CSPD had upregulated synchrony within several networks. At the baseline (v1) visit, CSPD showed exclusively increased connectivity between several dorsal attention and default mode ROIs, as well as between cerebellum and striatum. All upregulated connections were interhemispheric apart from ROI 12-ROI 20 (DM-DA). At follow-up (v2), CSPD had upregulated ipsilateral ROI-ROI connectivity between 3 VA and 1 SM ROI, as well as interhemispheric connections between single DA and SM as well as DM and STR ROIs. At follow-up, CSPD showed downregulated (interhemispheric) connections between SM ROI 66 and single DM and FP ROIs. Longitudinally (v2-v1), CSPD showed an increase in synchrony between STR-DM and CRB-VA; in addition, there was a relative decrease in synchrony between FP and SM as well as cerebellum ROIs, in addition to STR to VA.

**Table 3.**
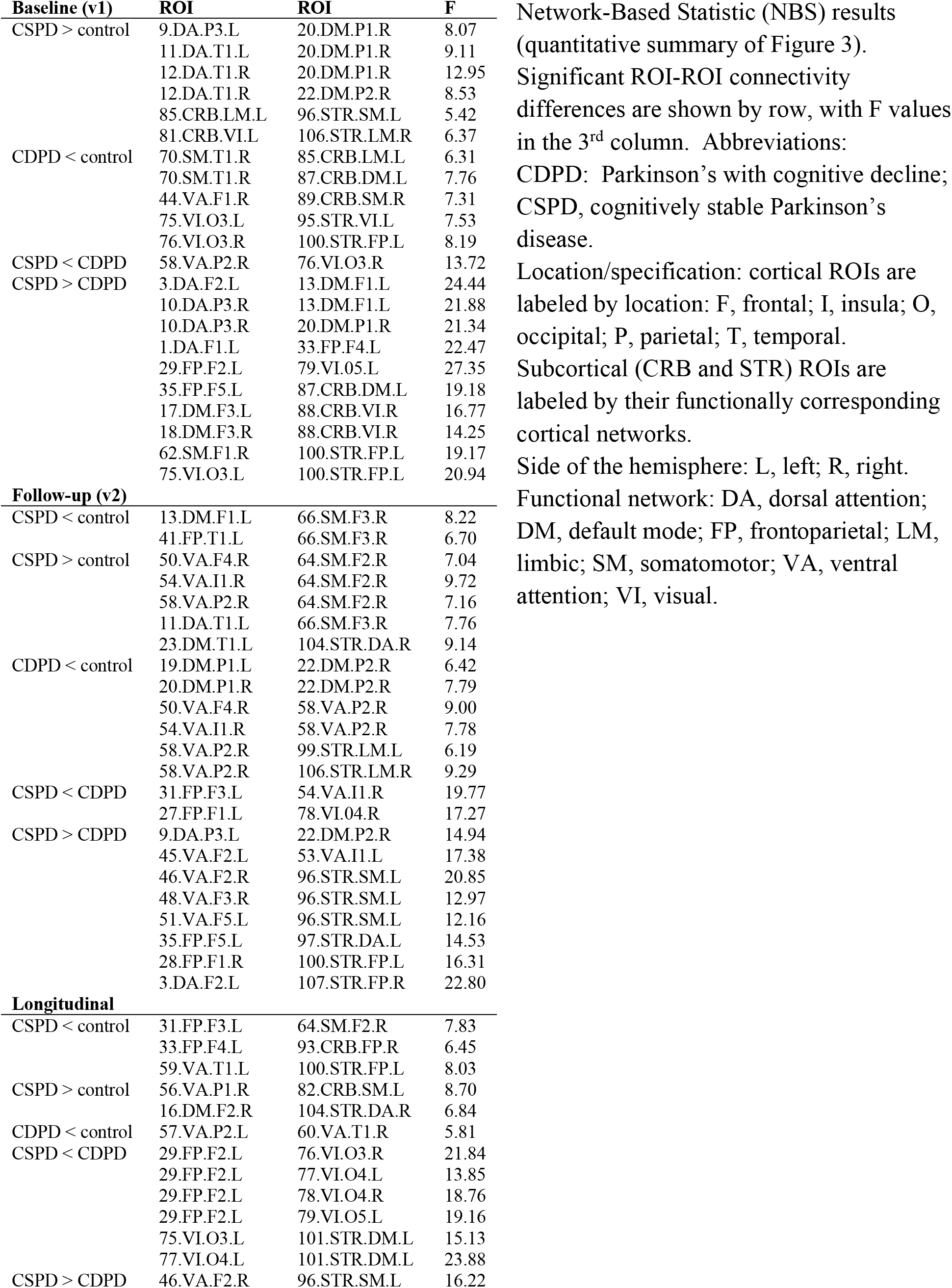
Network-based statistics (NBS) results

**Figure 3.**
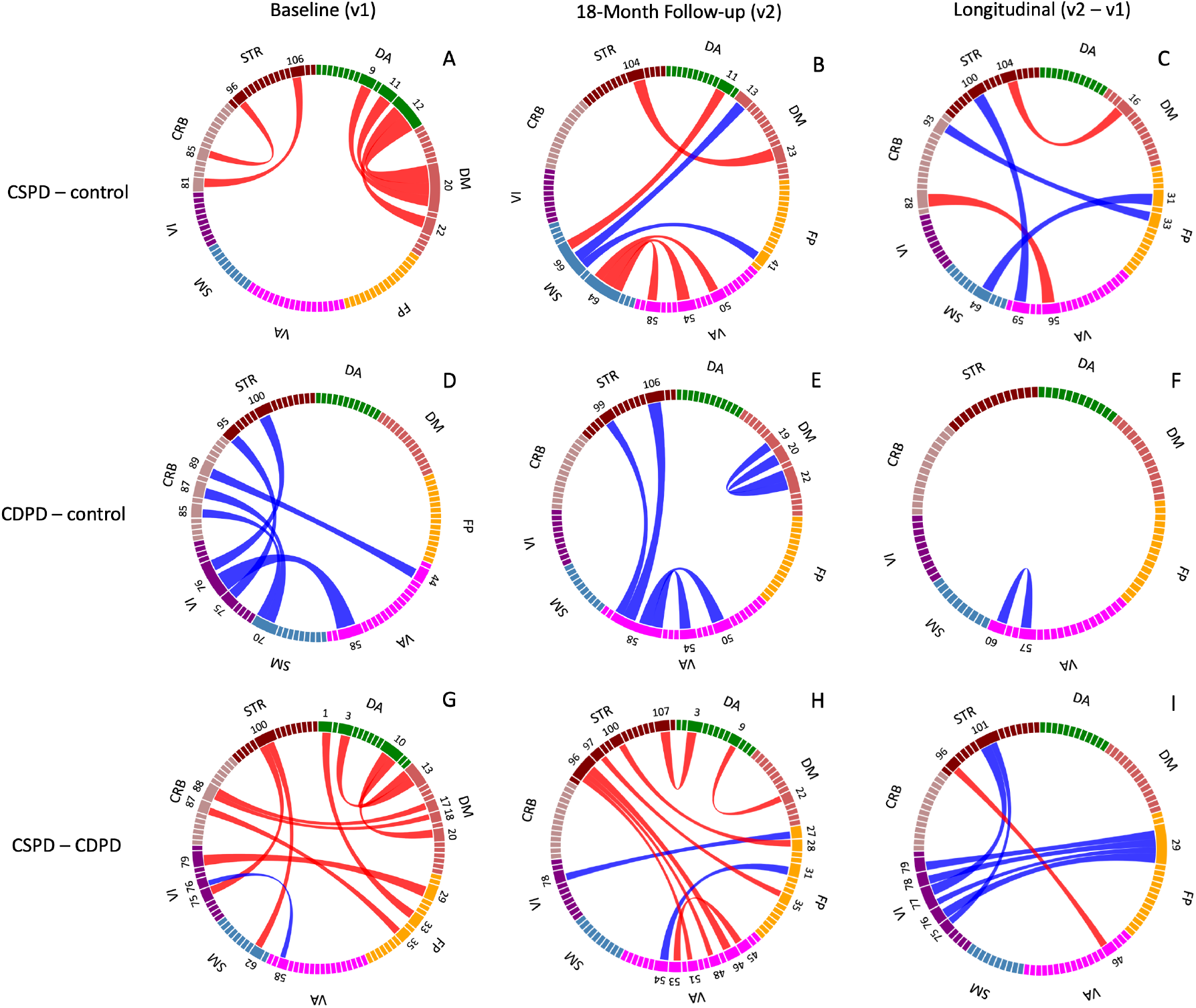
Network-Based Statistic (NBS) Results comparing control, cognitively stable PD (CSPD) and cognitive decline PD (CDPD) groups at baseline (v1), follow-up (v2) and change over 18 months (v2-v1). The width of the cords represents the relative significance of the results. Red cords and blue cords represent upregulated and downregulated networks, respectively, for each between-group comparison. Significant ROIs are labeled by their numeric indices only; anatomic locations can be found in Table 3. ***Abbreviations:*** DA, dorsal attention; DM, default mode; FP, frontoparietal; SM, somatomotor; VA, ventral attention; VI, visual.

In comparison to the control group, CDPD showed exclusively downregulated connections at both baseline and follow-up. At baseline, downregulated connections included striatum to VI, cerebellum to SM and VA, and V1 to VA. All were inter-hemispheric apart from bilateral STR-VI connections (75-95 and 76-100). At the follow-up (v2) visit, CDPD showed reduced intra-network synchrony for VA and DM, as well as inter-network, STR-VA. Several downregulated connections were within networks in the right hemisphere (DM, 20-22; VA 50-58 and 54-58). Comparison of v1 to v2 via subtraction showed little change between the visits, with a decrease in interhemispheric VA-VA connectivity only.

Not surprisingly, based on the comparisons of each PD group to control, CSPD demonstrated floridly upregulated connectivity compared to CDPD. CSPD had increased ROI-ROI connectivity in STR to SM and V1, CRB to DM and FP, DA to DM and FP, and FP-V1. At baseline, the only relatively downregulated connectivity was between one ROI each of the right V1 and VA networks (58-76). At the follow-up (v2) visit, CSPD showed upregulated ROI-ROI connectivity in intra-VA, STR to DA, FP, and VA, and DA-DM. At follow-up, the only downregulated interhemispheric connections were between 2 ROIs in the FP network and 1 ROI each in the V1 and VA networks. Longitudinally (v2-v1), CSPD showed a larger reduction in connectivity for multiple connections between left FP and V1 (both ipsi and contralateral), as well as STR ROI 101 and left V1 (75 and 77; ipsilateral), in comparison to CDPD. The only comparative increase in synchrony over time involved right STR-VA (96-46). Specific ROI-ROI relationships and corresponding F values for control versus CSPD versus CDPD differences are shown in Tables 3-4.

### Mean Inter- and Intra-Network Connectivity Analysis

Matrixes demonstrating the results of the mean inter- and intra-network connectivity analyses are shown in Figure 4. Significant results (*) that are similar to results of the NBS analysis are indicated by a dagger (†). At both baseline (v1) and follow-up (v2) time points, CSPD had greater mean inter-network connectivity than control in DA-DM, DA-SM, and VA-SM. Connectivity was increased at baseline in FP-CRB and STR-CRB. Several instances of reduced intra network connectivity were seen at either baseline or follow-up (VA-V1, SM-V1, and SM-CRB at baseline and DM-SM, FP-SM, and FP-V1 at follow-up), with SM intra-network down regulation also being seen at follow-up with NBS. Instances of significantly reduced intra-network connectivity were not observed in CSPD. These changes were borne out in the longitudinal results (v2-v1), with the addition of a comparative decrease in FP-VA, STR-VA, FP-CRB, and CRB-CRB and an increase in VA-V1 and SM-CRB connectivity.

**Figure 4.**
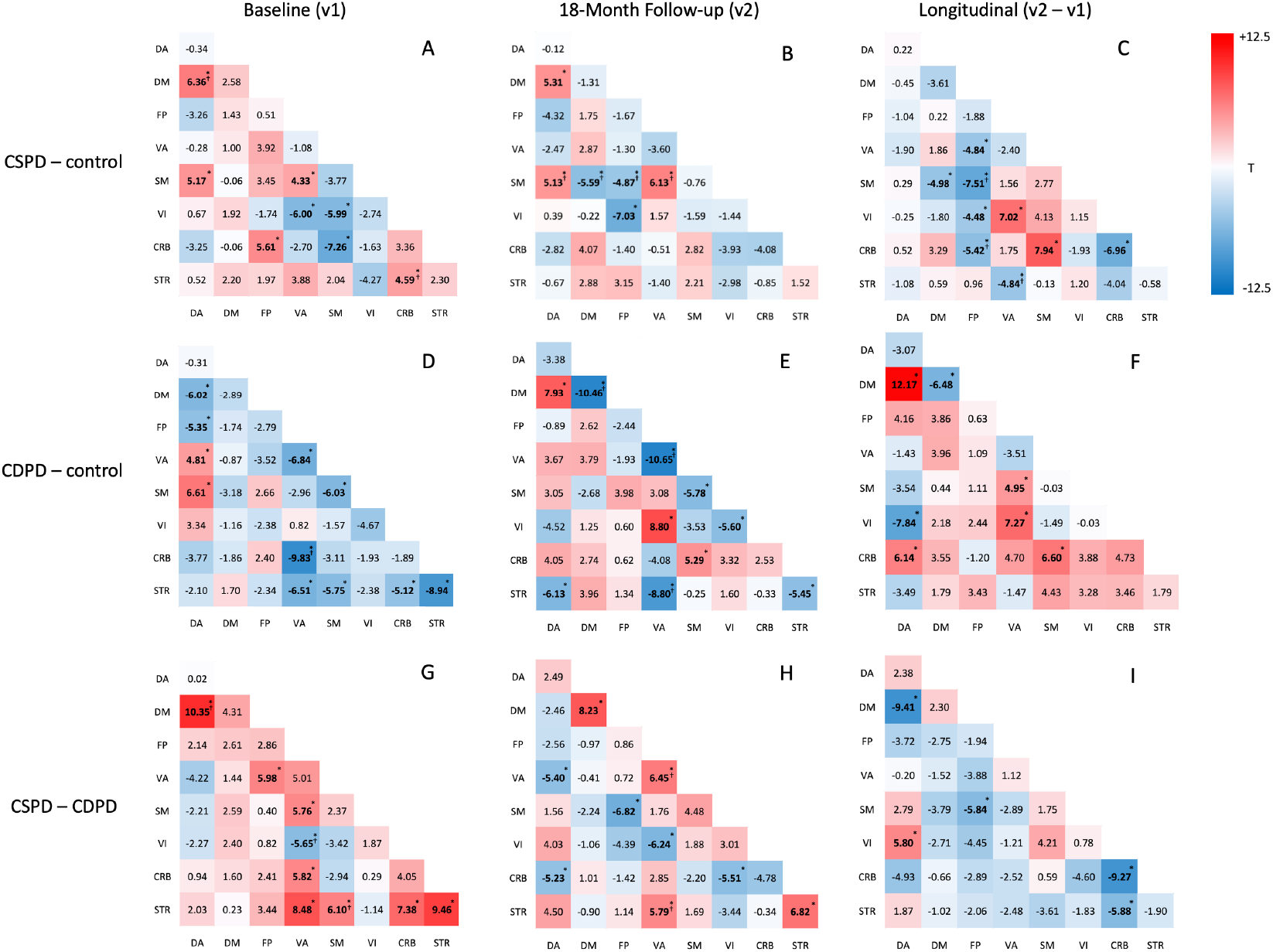
Mean intra- and inter-network correlations, compared between CSPD (n = 20), CDPD (n =10), and control (n = 35) R-fMRI, at baseline (v1), follow-up (v2), and longitudinal (v2-v1) time points. ROI-ROI connectivity differences for each two-group comparison were averaged within each network to yield mean intra-or inter-network correlations. Effect sizes (Welch’s t-test) for these comparisons are shown within each grid with * indicating significance after the Benjamini-Hochberg FDR correction for multiple comparisons at 0.05 level; ^†^Indicates overlapping results with the NBS analysis of ROI-ROI connectivity. **Abbreviations:** CDPD, cognitive declined Parkinson’s disease; CSPD, cognitive stable Parkinson’s disease; CRB, cerebellum; DA, dorsal attention; DM, default mode; FP, frontoparietal; SM, somatomotor; STR, striatum; VA, ventral attention; VI, visual.

In contrast to CSPD, CDPD showed several instances of reduced intra-network synchrony, involving DM (v2), VA-VA (both visits), SM-SM (both visits), V1-V1 (close to significant both visits), STR-STR (both visits). Reduced inter-network synchrony involved DA-DM, DA-FP, VA-STR, VA-CRB, SM-STR, and CRB-STR at baseline, and DA-STR and VA-STR at follow-up. Of the significant results at v1, decreased CRB-VA synchrony was also significant with NBS. At the follow-up (v2) visit, there was an increase in DA-DM connectivity (resembling the CSPD pattern), as well as VA-V1. Several instances of reduced within (DM-DM, VA-VA) and between (VA-STR) synchrony corresponded with the NBS results. A relative increase in between-network connectivity was observed in the longitudinal analysis, with the exception of connectivity decreases in DM-DM and DA-V1.

In comparison to CDPD, the CSPD showed generally higher intra-network connectivity for DM-DM (v2) and STR-STR (at both visits), as well as greater inter-network connectivity for DA-DM (v1), FP-VA (v1), SM and CRB-VA (v1), VA-STR (both visits), SM-STR (v1), and CRB-STR (v1) in CSPD. Only V1-VA connectivity was lower at baseline in CSPD compared with CDPD, a connectivity difference that persisted at follow-up. By the follow-up visit, differences in mean intra-network connectivity were less marked, with lower DA-CRB, DA-VA, FP-SM, VA-V1, and V1-CRB in CSPD. Of note, reduced DM-DM, VA-VA, and VA-STR synchrony in CSPD compared with CDPD was also seen with NBS at v2. Finally, the longitudinal (v2-v1) PD subgroup comparison showed relative decreases in connectivity in CSPD relative to CDPD for DA-DM, FP-SM, CRB-CRB, and CRB-STR.

Since CDPD (73.7 ± 6.8 yrs) was significantly older (p = 0.001) than CSPD (62.6 ± 8.8 yrs) at baseline (v1) and the mean intra-/inter-network functional connectivity in VI-DA, VA-SM, DM-SM, FP-STR, and STR-STR were correlated (p < 0.05) with age, the control subjects were resampled into two groups that resembled the age distributions of CSPD and CDPD, respectively. The mean intra- and inter-network comparisons between CSPD (n = 20) and their age-matched controls (n = 31), as well as between CDPD (n =10) and the age-matched controls (n = 18) at baseline (v1), follow-up (v2), and longitudinal (v2-v1) are shown in Supplemental Figure 3. These matrixes highly resemble those derived by comparing CSPD and CDPD to the entire control group, with the exception of slight changes in the degree of significance for connectivity differences within and between some networks. Therefore, we do not feel there is a large effect of age differences on the results.

## Discussion

Symptoms of Parkinson’s disease are caused by dysregulation of networks involving frontal-subcortical and frontal-cerebellar circuits (Alexander et al., 1986; Postuma & Dagher, 2006). Previous investigations using fluorodeoxyglucose or cerebral blood flow measures have shown motor and cognitive-related patterns involving regions of both increased and decreased blood flow or metabolism (Eidelberg, 2009). The motor-related pattern involves hypermetabolism in subcortical structures such as thalamus, globus pallidus, and pons, as well as primary motor cortex, with relatively reduced metabolism in the lateral premotor and posterior parietal regions (Peng et al., 2014). The cognitive pattern involves relative hypometabolism in the dorsolateral prefrontal cortex (part of the frontoparietal network in PINT), rostral supplementary motor area (Pre-SMA), precuneus (default mode), and posterior parietal regions, with increased metabolism in the cerebellum/dentate nucleus. Expression of the cognitive pattern is inversely correlated with verbal learning scores and increases with the degree of cognitive impairment in PD (Huang et al., 2007). While metabolism and resting state synchrony are not necessarily related measures, these metabolic investigations suggest we should expect to see both up- and down-regulated network activity in PD.

In the present study, we used PINT (Dickie et al., 2018) to optimize the location of nodes within cortical intrinsic connectivity networks. Because PD-related network dysfunction is known to affect the striatum and cerebellum, we also generated functionally defined regions within these structures for comparison to the PINT-defined cortical ROIs. We then compared patterns of inter- and intra-network connectivity between CSPD, CDPD, and control participants at baseline and 18-month follow-up. The most striking result from the NBS analysis was that PD participants who remained cognitively stable showed exclusively (at baseline) or predominantly (at follow-up) increased intra-network connectivity, whereas decliners showed exclusively reduced inter- and intra-(ventral attention and default mode) network connectivity, in comparison to the control group. In CDPD, mean intra-network (diagonal) connectivity was reduced for ventral attention, somatomotor, visual, and striatal networks, at both visits, and default mode network at follow-up. The decline group also had reduced intra-network connectivity of visual attention network, striatum, and cerebellum at both visits. Additional, less consistent decreases in intra-network connectivity were seen in both groups at either baseline or follow-up. Increased synchrony between the default-mode network and both the somatomotor and dorsal attention networks was observed in both CSPD and CDPD, suggesting that modulation of these networks may be a feature of PD that is not specific to cognitive decline (i.e. a “motor” pattern). Further, down-regulation of functional connectivity strength within and between multiple networks, including striatal intra-network connectivity, may be a feature of a “malignant” motor and cognitive phenotype (despite the lack of difference in UPDRS scores at baseline).

Multiple prior studies of PD have shown reduced functional connectivity within the default mode network, and between DMN and other cortical regions, as well as correlations between reduced DMN connectivity and the degree of cognitive impairment (Amboni et al., 2015; Baggio et al., 2015; Díez-Cirarda et al., 2018; Disbrow et al., 2014; Gorges et al., 2015; Hou et al., 2021; Tessitore et al., 2012; Wang et al., 2021; Wolters et al., 2019). Techniques used in these studies have varied, including seed-based, independent components analysis, and dynamic connectivity (to date, none have used PINT). In the present NBS analysis, CSPD had increased default mode inter-network connectivity in comparison to both controls and CDPD. Further, CDPD had reduced default mode to dorsal attention mean connectivity at baseline and decreased default mode intra-network connectivity at follow-up by NBS and mean calculations. Hence, findings of the present study suggest that decreased connectivity within the default mode network differentiates Parkinson’s patients who are destined to experience cognitive decline from those who will remain stable and supports the conclusion that reduced default mode network connectivity is associated with cognitive decline in PD.

Several previous studies evaluating R-fMRI in PD have found instances of increased functional connectivity in relation to cognition. Ruppert et al found increased default mode network connectivity in association with impairment of cognition in PD and PDD (Ruppert et al., 2021). Zahn et al. studied 9 each of healthy controls, PD with normal cognition, PD-MCI, and PDD, finding that intrinsic functional connectivity of the posterior cingulate cortex (PCC) to middle frontal gyrus, “middle temporal lobe”, precuneus, and cerebellum was greater in PD-MCI, and postulated that synchrony within these networks increases to compensate for cognitive decline (Zhan et al., 2018). Gorges et al, who studied 14 cognitively normal PD patients, 17 cognitively impaired PD patients, and 22 controls using a seed-based approach, found increased intrinsic functional connectivity “presenting as network expansions” in default-mode, bilateral frontoparietal cognitive control, ventral attention, motor, basal ganglia-thalamic, and brainstem intrinsic connectivity networks in cognitively unimpaired PD participants. In contrast, PD-cognitively impaired participants showed reduced extent of networks for default-mode, motor, and dorsal attention networks in comparison to controls (Gorges et al., 2015). This result is commensurate with results of the present study, which predominantly increased connectivity in CSPD and exclusively reduced connectivity CDPD by NBS and predominantly reduced mean intra-network connectivity in CDPD. Lack of involvement of the frontoparietal and dorsal attention networks in this NBS analysis is at odds with previous models of cognition in PD (Peng et al, 2014); however, the mean inter-network connectivity analysis did show increased connectivity between frontoparietal and cerebellar networks in CSPD, as well as reduced connectivity between default mode and dorsal attention networks in CDPD at baseline.

It is possible that R-fMRI differences reflect specific cognitive subtypes of PD. Devignes (Devignes et al., 2022) studied PD participants with frontostriatal (impaired attention, working memory and/or executive functions) versus posterior cortical (deficits in visuospatial function, episodic memory, and/or language) subtypes of MCI, demonstrating that patients with posterior cortical deficits had increased intra-network connectivity within the basal ganglia network compared with those with frontostriatal deficits. Frontostriatal MCI was accompanied by reduced inter-network connectivity between several networks (including default mode, sensorimotor, salience, dorsal attention, basal ganglia, and frontoparietal), in comparison to controls, unimpaired PD participants, and those with posterior cortical deficits. Since PD patients with the posterior cortical form of MCI are at higher risk to progress to dementia (Williams-Gray et al., 2013), this results seems to be at odds with the present study, but may reflect the subtypes included in our study (i.e. frontostriatal, with strong executive deficits).

One interesting observation from the present study is involvement of the ventral attention network in CDPD (reduced VA-VA, CRB-VA, STR-VA in comparison to control at both baseline and follow-up). Two other studies have described reduced VA functional connectivity in PD. One (Hou et al., 2021) reported reduced VA-VI inter-network, as well as reduced DM, V1, and SM intra-network connectivity in PD; another (Boon et al., 2020) found executive test performance to be positively correlated with DA-VA and negatively correlated with DA-SM connectivity in PD.

The role of the VA network in modulating responses to stimuli and susceptibility to visual hallucinations in Parkinsonism is a topic of recent interest. In PD patients, the onset of visual hallucinations frequently accompanies cognitive decline (and thus the transition to Lewy body dementia). Halluciations typically evolve from a sense of presence to visual illusions, then well-formed hallucinations of people and animals, suggesting that they are generated in part by misinterpretation of visual information rather than a perceptual loss. Current models of PD hallucinations hold an imbalance between the DM and DA networks leads to an ambiguous percent to which the VA network (driven in part by the amygdala) assigns incorrect salience (e.g. garden hose is perceived as a snake)(Shine et al., 2011). Although none of the participants in the present study suffered from visual hallucinations or dementia at baseline, it is possible that this connectivity pattern (reduced VA intra- and inter-network connectivity) is associated with cognitive vulnerability.

Few studies have evaluated resting state functional connectivity longitudinally in PD. One study (Filippi et al., 2021) compared connectivity between moderate-to-severe versus mild PD subtypes using a graphical analysis approach. They found that moderate-to-severe PD patients showed progressive alterations in global network properties (decreased mean nodal strength, local efficiency and clustering coefficient, and longer path length) relative to mild cases. Mild cases were characterized by increased connectivity, whereas moderate-to-severe cases showed progressive decreases in connectivity over time. In the present study, the NBS analysis a greater degree of change (with the development of decreased connectivity relative to controls) in CSPD versus CDPD, which showed persistently decreased connectivity and little change between the two visits. The finding echoes our findings with respect to structural connectivity, in which brainstem white matter microstructural integrity showed more rapid decline at earlier stages of disease (Pozorski et al., 2018). Since CDPD participants were not significantly impaired at baseline (but did have lower mean executive index compared with CSPD and control), this result suggests that the development of decreased network synchrony preceded clinically significant cognitive decline.

## Limitations

Ten of the 40 PD patients initially enrolled in this study dropped out prior to the second visit, largely due to disease progression with resulting difficulty attending study visits. Participant dropout reduced the size of our final longitudinal sample and may have biased the sample towards less impaired individuals. This sample size, in addition to not having collected dedicated visuospatial testing, limits our ability to comment on connectivity differences in PD cognitive subtypes. In addition, the participant groups were not ideally matched for age, probably in part because age is a risk factor for cognitive decline in PD. However, the results of the mean network connectivity analysis for age-matched subgroups (supplemental figure 3) yielded similar results to the entire sample. Nonetheless, this study yields intriguing data regarding R-fMRI in PD that extend those of previous studies by suggesting that connectivity profiles are associated with cognitive risk and should be investigated in larger independent samples.

## Data Availability

All data produced in the present work are contained in the manuscript

## Acknowledgments

This work was supported by Merit Review Award 101CX000555 (Gallagher, PI) from the US Department of Veterans Affairs Clinical Sciences R&D. Additional support for participant recruitment as well as data storage and analysis were provided by the University of Wisconsin Alzheimer’s Disease Research Center (NIH grant P50 AG033514; Asthana, PI). The authors declare no conflicts of interest with the work presented. The authors acknowledge Christopher Cox, PhD (Louisiana State University) for advice on the statistical design, and Rasmus Birn, PhD (University of Wisconsin, Madison), for advice on the data preprocessing. The data used in this study is considered property of the department of Veteran’s Affairs and is subject to VA data sharing policies. All participants provided written, informed consent prior to study participation under regulation of the University of Wisconsin Health Sciences IRB. This study is observational so not registered as a clinical trial. No materials from other sources are included in the work.

