## Supplementary Material for "A longitudinal evaluation of personalized intrinsic network topography and cognitive decline in Parkinson’s disease"

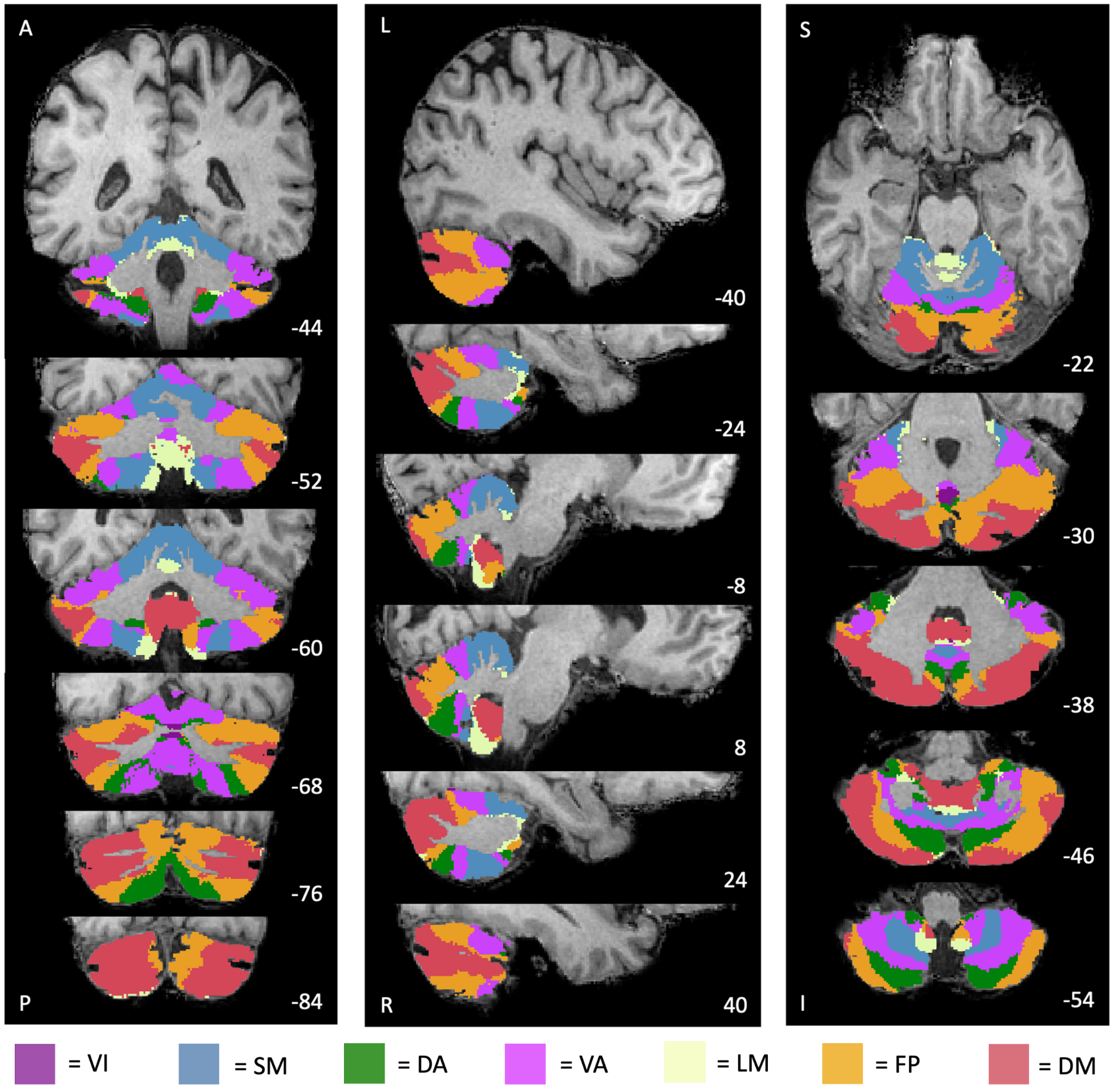


**Supplemental Figure 1.** Cerebellar regions of interest (ROIs) defined based on an intrinsic functional connectivity mask of the cerebellum (Buckner et al., 2011). Each cerebellar ROI corresponds to a cerebral network in Yeo's seven functional networks.
***Abbreviations:*** A, anterior; DA, dorsal attention; DM, default mode; FP, frontoparietal; I, inferior; L, left; LM, limbic; P, posterior; R, right; S, superior; SM, somatomotor; VA, ventral attention; VI, visual.

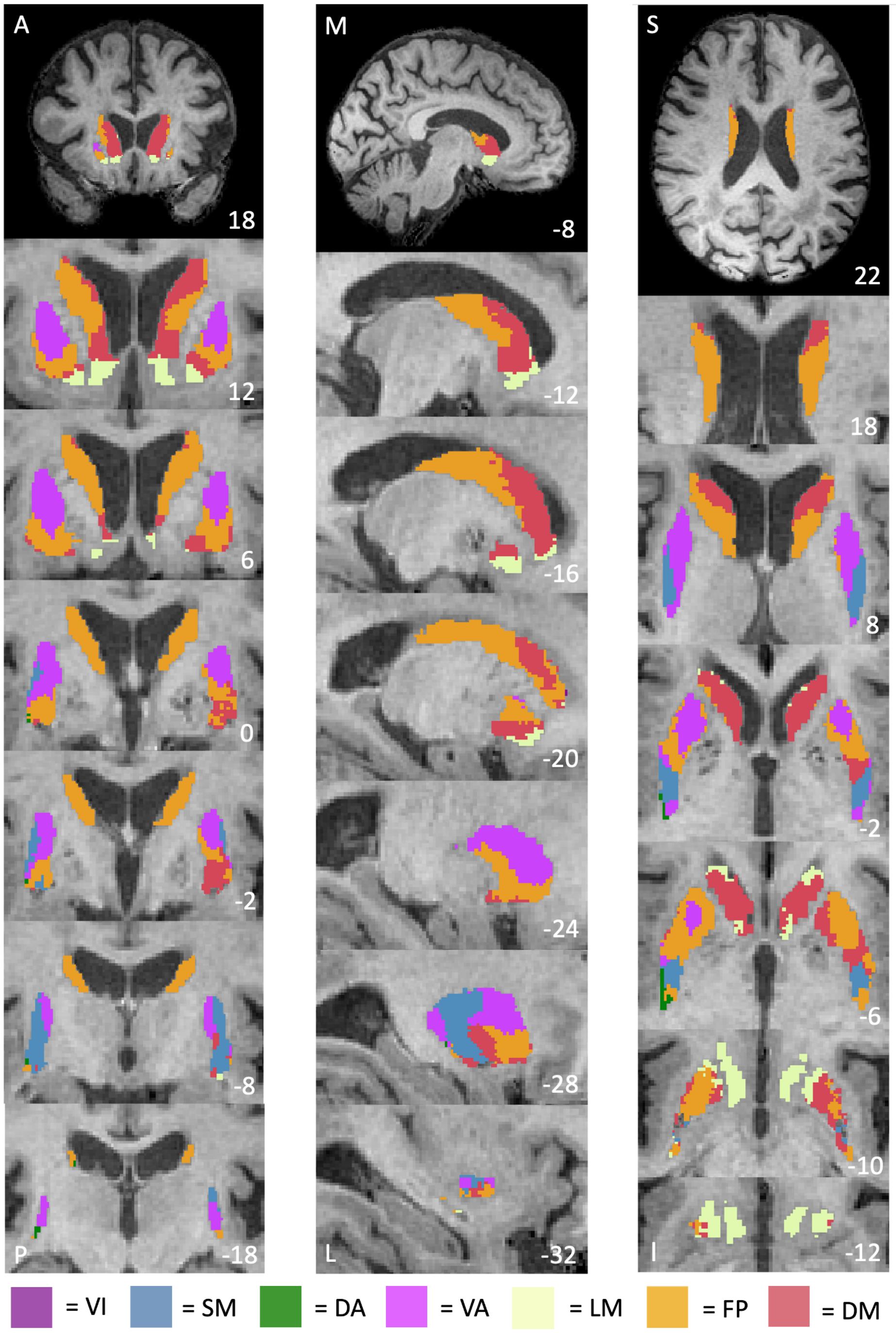


**Supplemental Figure 2.** Striatal regions of interest (ROIs) defined based on an intrinsic functional connectivity mask of the striatum (Choi et al., 2012). Each striatal ROI corresponds to a cerebral network in Yeo's seven functional networks.

***Abbreviations:*** A, anterior; DA, dorsal attention; DM, default mode; FP, frontoparietal; I, inferior; L, lateral; LM, limbic; M, medial; P, posterior; S, superior; SM, somatomotor; VA, ventral attention; VI, visual.

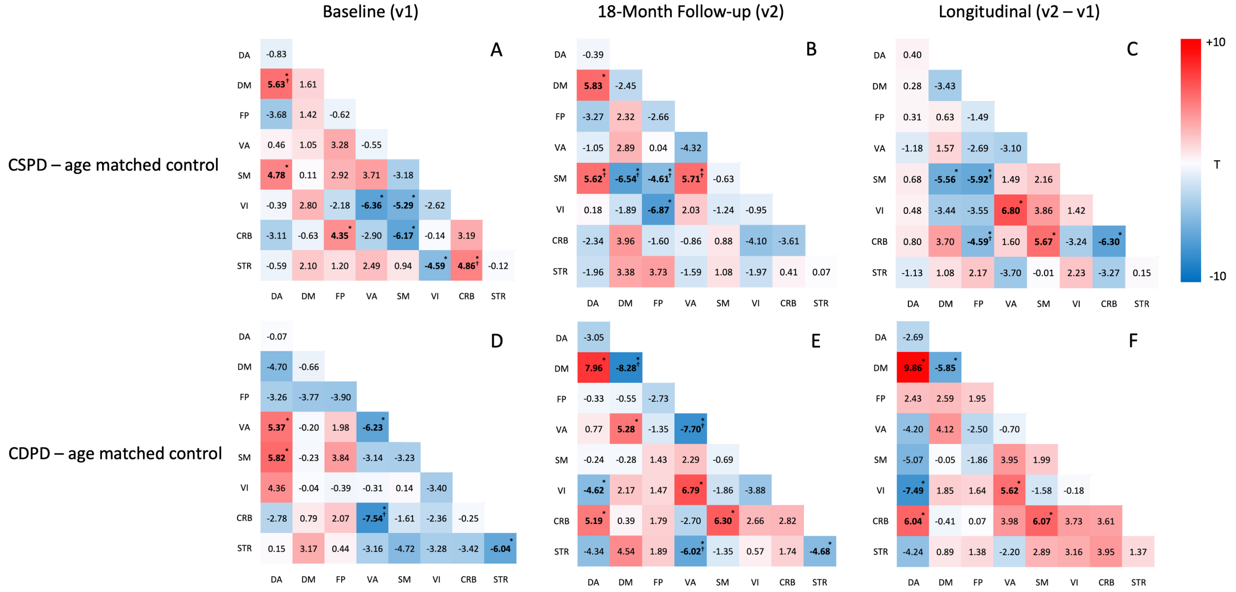


**Supplemental Figure 3.** Mean intra- and inter-network comparisons between CSPD (n = 20) and their age-matched controls (n = 31), as well as between CDPD (n =10) and the age-matched subgroup of controls (n = 18) at baseline (v1), follow-up (v2), and longitudinal (v2-v1). The ROI-ROI connectivity differences between two groups were averaged intra- or inter-network. The effect sizes are shown within each grid from Welch's t-test to compare each mean intra- or inter-network connectivity differences between groups,

*Indicates significance after the Benjamini-Hochberg FDR correction for multiple comparisons at 0.05 level.

†Indicates overlapping results with the NBS analysis of ROI-ROI connectivity.

**Abbreviations:** CDPD, cognitive declined Parkinson's disease; CSPD, cognitive stable Parkinson's disease; CRB, cerebellum; DA, dorsal attention; DM, default mode; FP, frontoparietal; SM, somatomotor; STR, striatum; VA, ventral attention; VI, visual.
